# Deconvoluting complex correlates of COVID19 severity with local ancestry inference and viral phylodynamics: Results of a multiomic pandemic tracking strategy

**DOI:** 10.1101/2021.08.04.21261547

**Authors:** Victoria N. Parikh, Alexander G. Ioannidis, David Jimenez-Morales, John E. Gorzynski, Hannah N. De Jong, Xiran Liu, Jonasel Roque, Victoria P. Cepeda-Espinoza, Kazutoyo Osoegawa, Chris Hughes, Shirley C. Sutton, Nathan Youlton, Ruchi Joshi, David Amar, Yosuke Tanigawa, Douglas Russo, Justin Wong, Jessie T. Lauzon, Jacob Edelson, Daniel Mas Montserrat, Yongchan Kwon, Simone Rubinacci, Olivier Delaneau, Lorenzo Cappello, Jaehee Kim, Massa J. Shoura, Archana N. Raja, Nathaniel Watson, Nathan Hammond, Elizabeth Spiteri, Kalyan C. Mallempati, Gonzalo Montero-Martín, Jeffrey Christle, Jennifer Kim, Anna Kirillova, Kinya Seo, Yong Huang, Chunli Zhao, Sonia Moreno-Grau, Steven G. Hershman, Karen P. Dalton, Jimmy Zhen, Jack Kamm, Karan D. Bhatt, Alina Isakova, Maurizio Morri, Thanmayi Ranganath, Catherine A. Blish, Angela J. Rogers, Kari Nadeau, Samuel Yang, Andra Blomkalns, Ruth O’Hara, Norma F. Neff, Christopher DeBoever, Sándor Szalma, Matthew T. Wheeler, Kyle Farh, Gary P. Schroth, Phil Febbo, Francis deSouza, Marcelo Fernandez-Vina, Amy Kistler, Julia Palacios, Benjamin A. Pinsky, Carlos D. Bustamante, Manuel A Rivas, Euan A. Ashley

**Affiliations:** Department of Medicine, Stanford University School of Medicine, Stanford, CA, USA; Department of Biomedical Data Science, Stanford University, Stanford, CA, USA; Institute for Computational and Mathematical Engineering, Stanford University; Department of Genetics, Stanford University School of Medicine, Stanford, CA, USA; Histocompatibility & Immunogenetics Laboratory, Stanford Blood Center, Stanford Health Care; Department of Statistics, Stanford University, Stanford, CA, USA; Department of Aeronautics and Astronautics, Stanford University, Stanford, CA; Department of Computational Biology and Swiss Institute of Bioinformatics, University of Lausanne, Lausanne, Switzerland; Department of Biology, Stanford University, Stanford, CA; Department of Pathology, Stanford University School of Medicine,, Stanford, CA; Medical Scientist Training Program, University of Pittsburgh and Carnegie Mellon University, Pittsburgh, PA; Chan Zuckerburg Biohub, San Francisco, CA; Department of Bioengineering, Stanford University, Stanford, CA; Sean N. Parker Center for Allergy and Asthma Research, Stanford University School of Medicine, Stanford, CA; Department of Emergency Medicine, Stanford University School of Medicine, Stanford, CA; Department of Psychiatry and Behavioral Sciences, Stanford University School of Medicine, Stanford, CA; Takeda Development Center, Americas, Inc; Illumina, Inc

**Author notes:** **Correspondence to:** Euan A. Ashley, MD PhD. Contributed equally (Victoria N. Parikh, Alexander G. Ioannidis, Carlos D. Bustamante, Manuel A Rivas, Euan A. Ashley).

## Abstract

The SARS-CoV-2 pandemic has differentially impacted populations of varied race, ethnicity and socioeconomic status. Admixture mapping and local ancestry inference represent powerful tools to examine genetic risk within multi-ancestry genomes independent of these confounding social constructs. Here, we leverage a pandemic tracking strategy in which we sequence viral and host genomes and transcriptomes from 1,327 nasopharyngeal swab residuals and integrate them with digital phenotypes from electronic health records. We demonstrate over-representation of individuals possessing Oceanian and Indigenous American ancestry in SARS-CoV-2 positive populations. Genome-wide-association disaggregated by admixture mapping reveals regions of chromosomes 5 and 14 associated with COVID19 severity within African and Oceanic local ancestries, respectively, independent of overall ancestry fraction. Phylodynamic tracking of consensus viral genomes reveals no association with disease severity or inferred ancestry. We further present summary data from a multi-omic investigation of human-leukocyte-antigen (HLA) typing, nasopharyngeal microbiome and human transcriptomics that reveal metagenomic and HLA associations with severe COVID19 infection. This work demonstrates the power of multi-omic pandemic tracking and genomic analyses to reveal distinct epidemiologic, genetic and biological associations for those at the highest risk.

## MAIN

Two central questions from the COVID-19 pandemic remain unresolved: who is at risk of severe disease, and why? Genome-wide-association-studies (GWAS) from the COVID-19 Host Genetics Initiative and others have identified up to 13 genetic loci associated with COVID19 infection, hospitalization and critical illness.^1,2,3^ Among these loci is a chromosome 3 haplotype that entered the human population from Neanderthals, suggesting that genetic ancestry plays an important role in susceptibility and severity in SARS-CoV-2 infection.^4^ At the same time, epidemiologic studies have shown that comorbidities, sex and socioeconomic status are strongly associated with infection prevalence and disease severity.^5–8^ Several groups have reported higher incidence of COVID19 infection and higher disease severity among Hispanic/Latino and African American racial and ethnic groups.^5,7^ Because social constructs of race and ethnicity covary with ancestry-related genetic markers, such associations may confound the study of COVID19 host genetic susceptibility. Genetic ancestry inference along the genome represents a powerful tool to examine genetic risk within the context of multi-ancestry genomes. These analyses are independent of the socioeconomic factors associated with race and ethnicity, due to the random shuffling of genomic ancestry segments, even between siblings, via recombination. Further, viral phylodynamics, host immunity (e.g., HLA typing), and metagenomics are important potential contributors to disease severity that can vary with ancestry and social factors and can be clarified with this omics-based approach.

To build the complex database necessary for these investigations, residual viral transport media (VTM) from SARS-CoV-2 clinical diagnostic tests were randomly prospectively collected from March 2020 to July 2020 and linked to structured clinical information from the electronic health record. 938 SARS-CoV-2 positive and 389 SARS-CoV-2 negative NP swab residuals were sequenced from a total of 892 (571 positive and 321 negative) patients (**Figure 1A&B**). Host whole genome sequencing was aligned and called using methods for low pass data ^9–11^(mean of mean coverages: 2.56X±2.50X(SD), **Figures 1C**), followed by phasing and imputation with GLIMPSE **(Figure S1)**.^12^ Shotgun RNAseq of initial samples yielded high coverage of much of the viral genome for samples with clinical test CT values <35 regardless of RNA yield, which improved with primer-based capture (**Figure 1D** and **S1D**). For digital phenotype abstraction, we generated a COVID19 clinical severity score based on the ordinal scale proposed by the World Health Organization (**Table S1**). Clinical data were obtained through the STAnford Research Repository (STARR), specifically the STRIDE data management system, which is populated from patients’ clinical and biospecimen data.^13^ Severity score was calculated on the date of sample collection and daily for one month before and indefinitely after (**Figures 1E&F** and **S2**).

**Figure 1.**
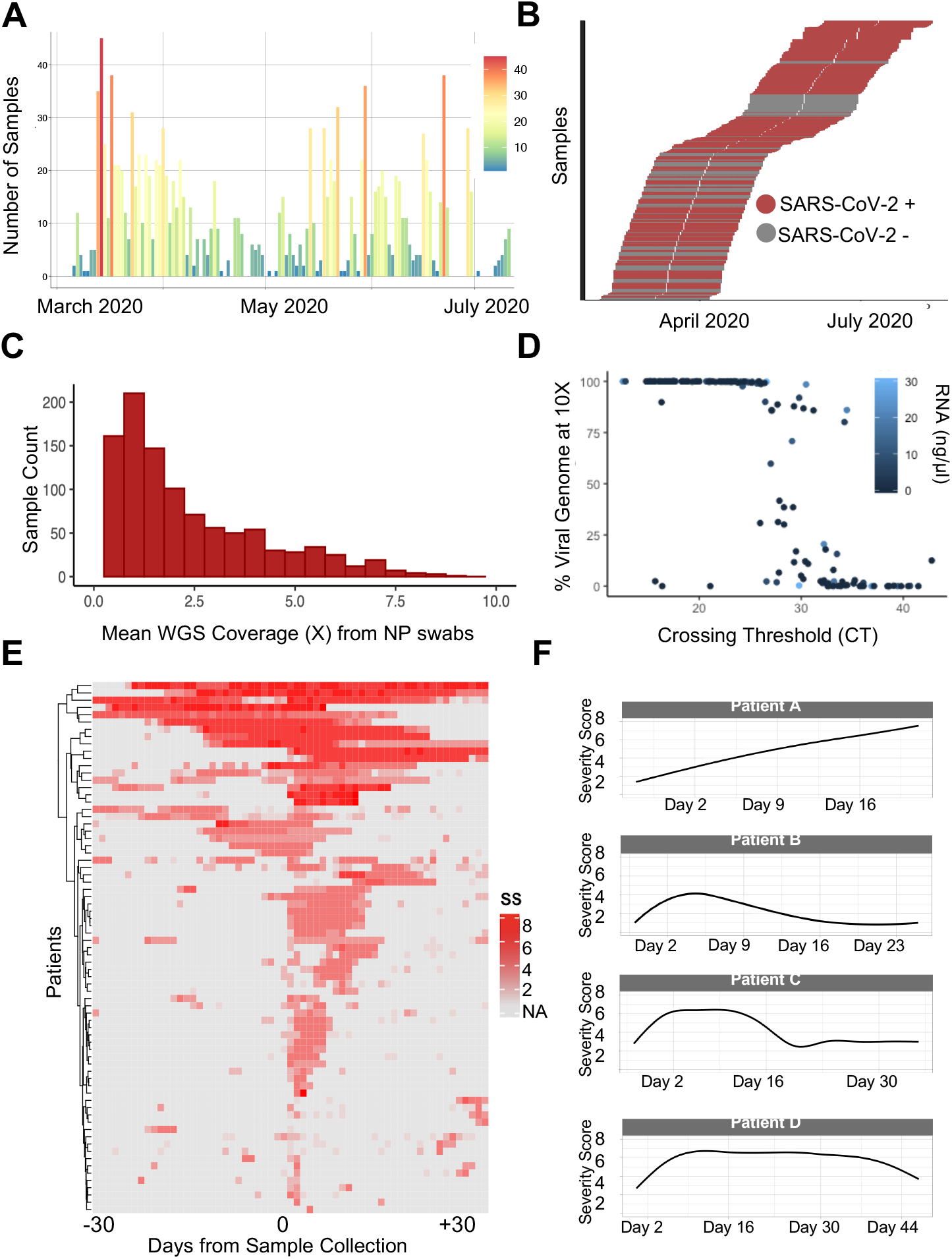
SARS-CoV-2 Pandemic tracking from residual NP swabs and abstracted EHR data. **(A)** We collected 1327 NP swab residuals from a single Northern California Hospital System between March and July 2020. Colors represent the number of samples collected over time (x-axis, see legend). **(B)** These included 938 SARS-CoV-2 positive and 389 SARS-CoV-2 negative swabs that were collected in parallel during this period. Time over which clinical data was abstracted for each individual is shown (30 days before and after sample collection date.) **(C)** Whole genome sequencing from DNA isolated from 150 ul of NP swab VTM yielded sequence on >95% of samples with mean of means coverage 2.6X. **(D)** RNA sequencing using shotgun sequencing recovered consensus SARS-CoV-2 sequence on the majority of NP swabs with a clinical PCR CT value <30. ARTIC primer enrichment increased this yield **(Figure S1D). (E and F)** Severity scores abstracted directly from the electronic health record daily for thirty days before and after the positive NP swab test on all included patients with severity score ≥ 4 (hospitalized, needs oxygen) demonstrates significant variability in patient course.

We used genetic ancestry inference to identify subpopulations highly impacted by the COVID19 pandemic. Self-reported race and ethnicity re-demonstrates prior reports of over-representation of minority race and ethnic populations within the US amongst COVID19 positive patients (**Figure S3**). Based on genetic ancestry inference, a higher proportion of individuals of Oceanian (Pacific Islander) and Indigenous American genetic ancestry exists in COVID19 cases as compared to negative controls (**Fig 2A**, 44% vs. 26% for individuals having Indigenous American genetic ancestry, associated with Hispanic populations (χ^2^ = 99, p<1e-10), and 4.6% vs. 2.3% for individuals having Oceanian ancestry, associated with Pacific Islander origins (χ^2^ = 13.9, p=2e-4)). Evaluation of temporal trends of genetic ancestry in case positivity reveals early predominance of European ancestry followed by a significant increase in Indigenous American ancestry after May 2020. These findings are recapitulated by self-reported ethnicity, as the majority of COVID19 cases between May and July 2020 self-identified as Hispanic/Latino in the medical record, an ethnicity often associated with mixed Indigenous American and European genetic ancestry (**Fig 2B**).

**Figure 2.**
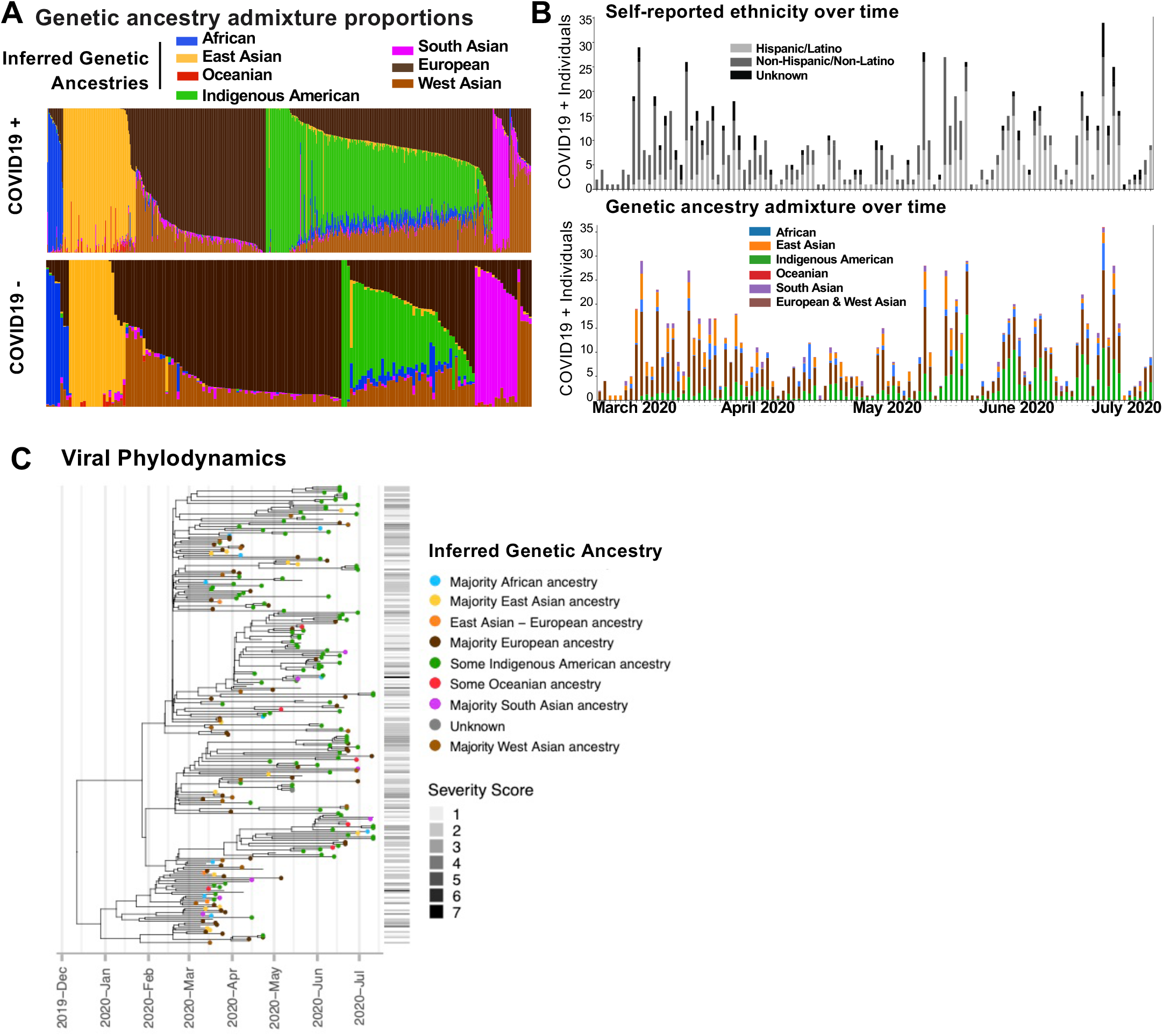
Genetic ancestry inference allows identification of high risk populations and tracing of transmission over time through its interaction with viral phylogeny and disease severity. **(A)** Genetic ancestry admixture of individuals with positive versus negative COVID19 tests in the present study. Individuals with Indigenous American ancestry are overrepresented in cases, whereas controls show more European and South Asian genetic ancestry. **(B)** Self-reported (top) and genetic ancestry (bottom) of enrolled COVID19+ individuals over time reveals disproportionate representation of Hispanic/Latino ethnicity and Indigenous American ancestries during summer pandemic wave, whereas the first wave is seen to have predominantly affected non-Hispanic individuals and individuals of European genetic ancestry. **(C)** Phylogenetic reconstruction of SARS-CoV-2 sequences. Tip colors correspond to the inferred genetic ancestry of the infected hosts, whose consensus SARS-CoV-2 sequences were isolated and used for inferring the viral phylogeny. Horizontal lines at the bottom of the phylogeny indicate host severity scores corresponding to the tips of the phylogeny. Severity score codes are displayed in Table S1.

We also investigated the role of viral phylodynamics in COVID19 severity and its potential interaction with individual ancestry and disease severity. Consensus viral genomes (10X coverage at >99% of the genome) were recovered for 255 samples from unique, unrelated individuals. The estimated time to the most recent common ancestor of observed samples is December 11, 2019 with a 95% Bayesian CI of (2019-10-27, 2020-01-12). However, the phylogenetic reconstruction (**Figure 2C**) reveals an early introduction in the area between late December 2019 and early January of 2020 with several independent introductions later in February 2020. While about 37% of the infected individuals in the sample have Indigenous American ancestry, there is no evidence of exclusive transmission amongst individuals of this ancestry. Other genetic ancestries also do not sort with clades, though a single clade from early in the pandemic had fewer Hispanic individuals, consistent with the first wave prior to May 2020, in which European genetic ancestry individuals were enriched (**Figure 2C**). We also tested the hypothesis of association between viral lineages and disease severity. No association was found between specific clades and severity score at the time of NP swab (**Figure 2C**).

After adjusting for age, sex and BMI (known correlates of disease severity^14–16^) together with overall genetic ancestry proportion, we assessed association of genetic ancestry at a given genomic position with the COVID19 severity score as an ordinal outcome (admixture mapping). Because our captured case population was enriched for non-European ancestry groups, we were able to perform admixture mapping for six ancestries (African, Native American, Oceanian, South Asian, East Asian, and European/West Asian). This analysis revealed loci in chromosomal regions of African and Oceanic ancestry that met genome-wide significance, determined as previously described by Shriner et al.^17^ (**Figure 3**). In segments of African ancestry, a region of chromosome 5 (5:34675449-35163719, GRCh38.p13) was associated with increased disease severity (p=0.03 after multiple test correction), while in segments of Oceanian ancestry, a region of chromosome 14 (14:44073392-44149995) was also associated with high severity COVID19 (p=1.2e-07 after correction). Two SNPs in these regions (rs13167486 and rs4608241 respectively) were reported in the same genome-wide interaction analysis of pathological hallmarks of Alzheimer’s Disease^18^ and map to two long noncoding (lnc)RNAs, SPRY4-AS1 and LINC02307. These lncRNAs are predicted to bind a set of miRNAs^19^ whose empirically confirmed target genes are enriched in brain, as well as epithelium (e.g., lung).^20, 21^ LncRNAs have an important role in antiviral immunity both via their effect in immune cells as well as direct interaction with the viral genome.^22,23^

**Figure 3.**
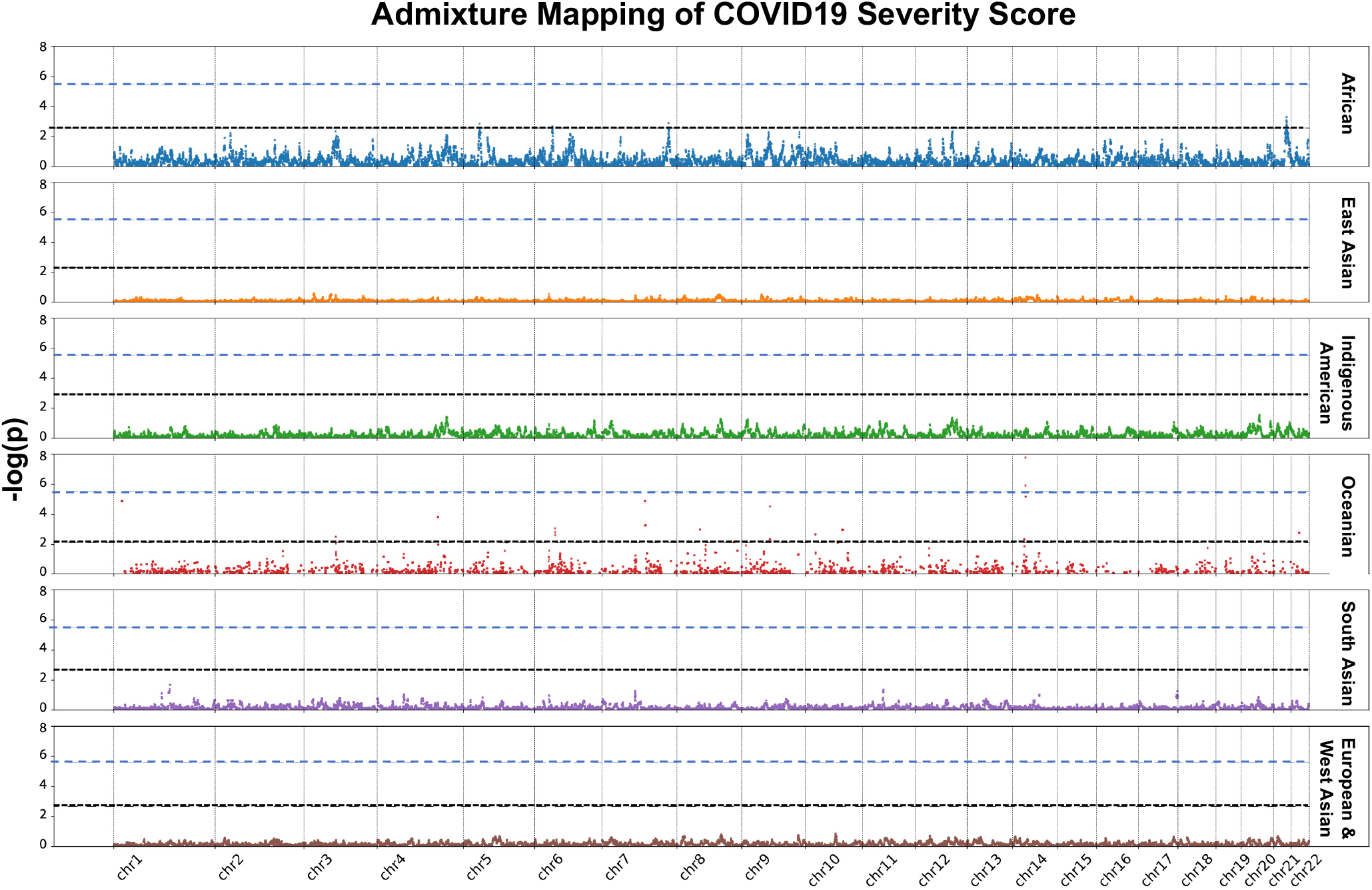
Admixture mapping shows association of ancestry along the genomes of COVID+ patients with severity score. Ancestry-specific risk loci found in Chromosome 5 and 14 in African and Oceanian ancestries, respectively, were found to be associated with the same functional mechanism. Genetic ancestry associations were first corrected for overall ancestry proportions of patients, for BMI, and for age. Lower black dotted-lines indicate less stringent ancestry-specific multiple test correction for a p-value of 0.05, using the method of Shriner et al., while higher, more stringent blue dashed lines indicate naive Bonferroni corrections for the same.

We next established a resource of summary statistics for COVID19 severity versus host genotype by genetic ancestry, host HLA type, and metagenomic alignments (https://covid-omics.org/results), and have contributed host genetic summary data and viral consensus sequences to the COVID-19 Host Genetics Initiative^3^ and GISAID,^24^ respectively (**Figure 4A**). Using this resource, we explored the potential contribution of the nasopharyngeal microbiome and HLA-type as biological determinants of COVID19 severity. A UMAP plot of microbiome species abundance shows clustering largely independent of severity (**Figure 4B**); however, a regression of species abundance against COVID19 severity (controlling again for age and BMI) revealed enrichment of *Paracoccus yeei* transcripts in high severity cases, a bacterium that causes opportunistic infections in critically ill,^25^ organ transplant,^26^ and dialysis patients, ^27,28^ indicating an association with immune compromise and severe illness (**Figure 4C**, p =7e-04 after Bonferroni correction). The HLA-B*07:02 allele (common prototype allele for the serotype B7) was associated with elevated risk of high severity score (OR 2.7 [1.4, 5.1], p=2.9e-03), whereas the HLA-C*15:02 allele (common prototype allele for the serotype Cw15) was associated with risk reduction (OR 0.12 [0.02, 0.82], p=1.41e-02) (**Figure 4D**). HLA-B*07:02 presents epitopes from the SARS-CoV-2 *N* gene and *Orf1ab*,^29^ and the HLA-C*15:02 allele contains two distinctive amino acid substitutions at residues 113 and 116 located within the peptide binding groove. HLA-C*15:02 was associated with milder disease in the first SARS epidemic,^30^ and is predicted to bind a SARS-CoV-2 Spike protein epitope.^31^

**Figure 4.**
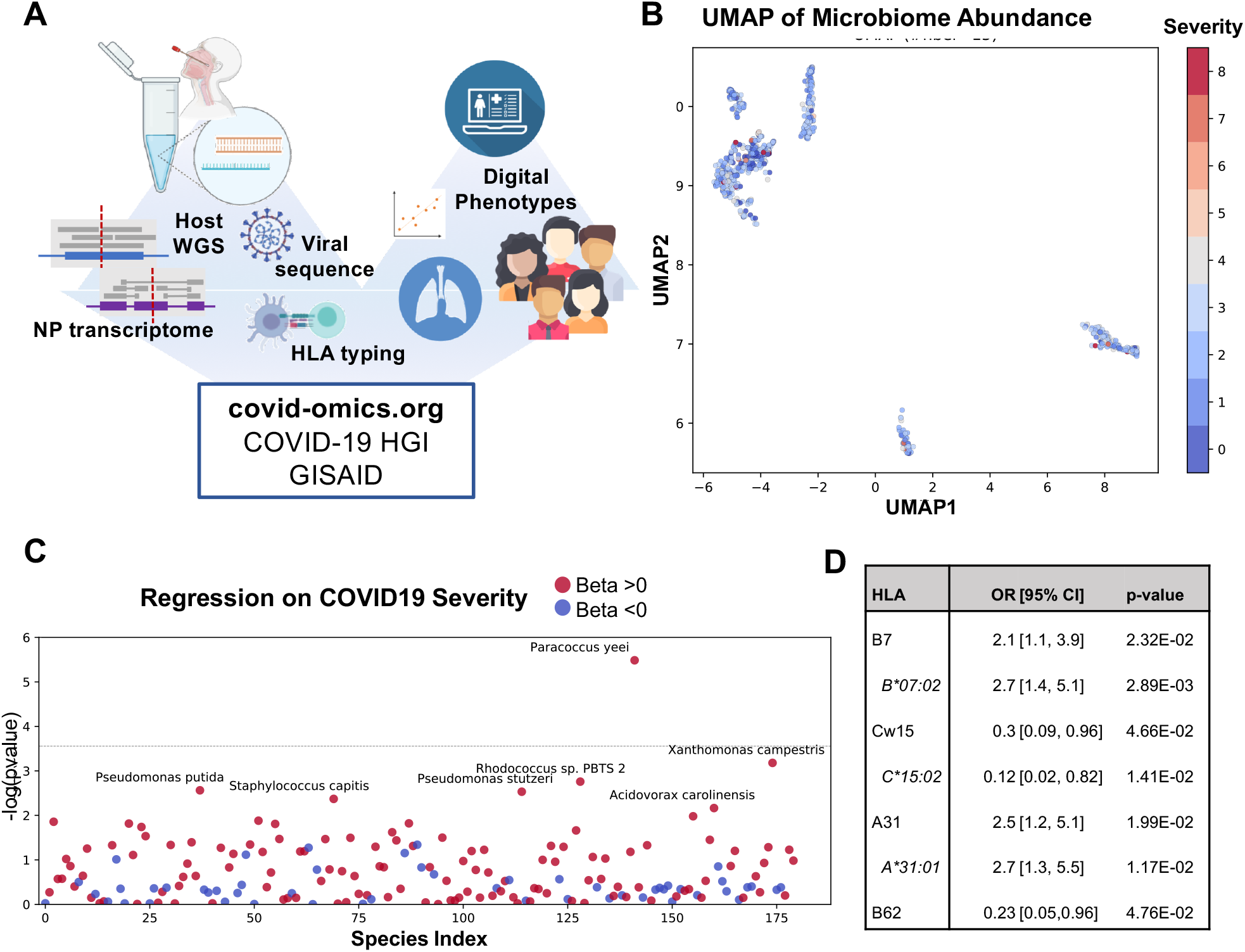
**(A)** Figure representing data resource. **(B)** Uniform manifold approximation and projection (UMAP) of Nasal Microbiome abundance colored by severity score. **(C)** Regression of species-specific transcript abundance against continuous disease severity, corrected for BMI and age, identified *P. yeei* abundance in the nasopharyngeal microbiome as associated with high severity COVID19 infections (Bonferroni adjusted p=7e-04). (**D**) HLA allele and serotype (italics) association with COVID19 severity after ancestry adjustment by Cochran Mantel Haenszel test.

These results represent a substantial effort to assemble host and viral genomic, transcriptomic and digital clinical data from a diverse cross-section of the socioeconomic, racial and ethnic groups hit hardest by the COVID19 pandemic. We show that ancestry inference can be used to track changes in the affected population in real-time, demonstrating that Hispanic/Latino groups (associated with Indigenous American genetic ancestry) were disproportionately affected during a second pandemic wave. This is consistent with the model that this second wave was driven not by introduction from travelers (likely the source of the first wave), but by economic pressure on essential service workers to leave their homes and family units, enabling viral spread.^32^ Phylodynamic overlay on this at-risk population further supports this conclusion, demonstrating that viral clades did not differentially affect ancestral groups, nor did they confer differential disease severity during six months of prospective enrollment. Thus, the impact of introduction of viral variants on community spread was likely less than that of exposure related to essential services work.

In addition to the use of ancestry inference to track the impact of the pandemic on ancestral sub-populations, genomic regions associated with COVID19 severity in the context of local African and Oceanian ancestries highlight potentially novel pathobiology. Both regions are near lncRNAs, non-coding transcripts that can interact with other small noncoding RNAs to alter viral immunity and host gene expression, one of which is expressed in lung epithelium, consistent with potential biological significance to SARS-CoV-2 infection.^19,21^ Admixture mapping and local ancestry disaggregation were necessary to reveal these markers, which would likely otherwise be masked by social and economic determinants of severity that disproportionately affect these ancestral populations.

As the populations at highest risk of severe outcomes in a pandemic (due in large part to health and socioeconomic inequities) are not proportionally represented in existing datasets, the development of our real-time data collection strategy from clinical swab residuals was critical to assessing the relevance of ancestry-specific genetic variation.^33^ We present a resource combining summary host and viral genomic, metagenomic and transcriptomic data with highly evolved digital phenotype abstraction from extant EHR data to help deconvolve genetic environmental and social factors while tracking spread across the community. This system can be applied in real-time to model individual and population trajectories in future emerging global infectious diseases (https://covid-omics.org/results), and can help illuminate underlying biology and medical insights relevant to diverse populations that can be otherwise missed or ignored due to confounding with the social and economic factors that are also associated with race and ethnicity in diverse populations.

## METHODS

### Sample Collection and diagnostics

Residual VTM from SARS-CoV-2 positive nasopharyngeal swabs collected during clinical assessment of asymptomatic and symptomatic patients at Stanford Healthcare were used in accordance with the Stanford School of Medicine Institutional Review Board. RT-qPCR targeting the *envelope* gene or ORF1ab were used to detect infection. 152 samples with detectable SARS-CoV-2 RNA and 8 with undetectable virus by RT-qPCR were included. Positive samples were defined as those crossing threshold (CT) of 40 cycles or less on the RT-qPCR or positive Transcription-Mediated Amplification (TMA) diagnostic tests used at Stanford Health Care clinical laboratory.^34^

### EHR Data Abstraction and Severity Score Development

A critical task is to determine for every sampled patient the disease severity from the Electronic Health Records (EHR). To accomplish this task, we used as the “COVID-19 Clinical Severity Scale” an adapted version of the “Ordinal Scale for Clinical Improvement” proposed by the World Health Organization in the COVID-19 Therapeutic Trial Synopsis (Draft February 18, 2020, Table S1). This scale categorized the COVID-19 severity according to the level of care and oxygen support. Scores 1 to 2 include patients not requiring supplemental oxygen support or hospitalization. However, the WHO definition of these scores were modified due to their vague scope. Thus, score “1”, originally described as “no limitation of activities”, was modified to “asymptomatic patient”, and score “2”, from “limitation of activities’’ to “symptomatic patient” (symptoms were extracted and curated from EHR billed diagnoses). Scores 3 to 4 include patients hospitalized, with score 4 assigned only to those requiring non-invasive supplemental oxygen (oxygen mask). Scores 5 to 7 are defined as “severe disease” based on level of oxygen support. Thus, “score 5” is for patients requiring high flow oxygen and “score 6” mechanical ventilation. “Score 7” includes critically ill patients requiring, in addition to ventilation, the administration of specific medications (pressors), dialysis, or extracorporeal membrane ventilation (ECMO). A custom algorithm was written that abstracted digital phenotypes from each chart (see Table S1, “EHR annotations”). First, SARS-CoV-2 positive status was confirmed based on clinical test reports abstracted from the EHR. SARS-CoV-2 negative patients were assigned a score of zero. For SARS-CoV-2 positive patients, starting with the highest score (8, death) and working down, if criteria were met, the individual was assigned that score. If no clinical notes were available for data abstraction, then a severity score was not assigned and these individuals were not included in severity score based analyses. We calculated the score for any given date and assigned the maximum value according to the EHR annotations defined for every score (Supplementary Table 1). For example, patients with annotations for both ventilation and the administration of pressors received a score “7” for that day. Clinical data were obtained through the STAnford Research Repository (STARR), a Stanford Medicine’s approved resource for working with clinical data for research purposes extracted from the Epic database management system used by the Stanford hospitals. Specifically, we queried the STRIDE data management system, which is populated from patients’ observational clinical, research, and biospecimen data^13^.

### Nucleic Acid Extraction

DNA; host genomic DNA was extracted from 200ul of VTM inoculated with nasopharyngeal swabs. Using a modified Qiagen DNEASY blood and tissue kit protocol and quantified using fluorometric readings (Protocols.io doi: dx.doi.org/10.17504/protocols.io.bi8xkhxn). Total RNA was extracted from 200ul of VTM using a modified Ambion MirVana mRNA kit protocol (Protocols.io doi: dx.doi.org/10.17504/protocols.io.bi8ykhxw) or Zymo Research Quick-Viral RNA extraction kits (R1041) and quantified using fluorometric readings.

### Host gDNA library preparation and sequencing

Using 1-10ng of host gDNA, the Illumina Nextera Flex library preparation was performed according to manufacturer’s protocol (Protocols.io doi: dx.doi.org/10.17504/protocols.io.bi8zkhx6). To allow for multiplexing, gDNA was barcoded using IDT-ILMN Nextera DNA UD Indices, a set of 10bp index adapters from Illumina. Indexed samples were diluted to 4nM, pooled, and analyzed on an Agilent TapeStation to ensure the mean DNA fragment size was ∼300bp. Pooling and library quality was further assessed by sequencing the pool using a V3 MiSeq flow cell. 160 samples were pooled and sequenced for 76 cycles, paired end reads. For the purpose of QC, ∼ 50 million reads were obtained and Q30 was determined to be >92%. If needed the pool was normalized (balanced) to ensure equal representation of each sample. The library was then sequenced on an Illumina NovaSeq 6000 using an S4 300 cycle flow cell.

### Viral RNA library preparation and sequencing

After extraction, RNA acquired from 100 ul nasal swab media were incubated with recombinant RNAse-free DNase (Qiagen, Inc.) per manufacturer’s instructions for 15 minutes, followed by SPRI bead (GE Healthcare) purification to remove residual DNA remaining in each sample. A fixed volume (5uL) of the resulting RNA from each sample, together with a fixed mass (25pg) of the External RNA Controls Consortium RNA spike-in mix (ERCC RNA spike-in mix, Thermo Fisher), served as input for SARS-CoV-2 metatranscriptomic next generation sequencing (mNGS) library preparation (dx.doi.org/10.17504/protocols.io.beshjeb6; a modification of Deng et al. (2020)^35^).

For samples collected after May 2020, SARS-CoV-2 ARTIC V3 amplicon libraries were made from extracted total nucleic acid for whole genome sequencing using previously reported protocols ^[1]^. Briefly, 3 ul of total nucleic acid was used as input for a randomly primed cDNA synthesis reaction. This cDNA served as input for 30 cycles of amplification with ARTIC V3 primers (github repo: artic-network/artic-ncov2019), and was then diluted 1:100 before tagmentation. Adaptor tagmentation was performed using homebrew Tn5, and 8 cycles of index PCR was performed using unique dual barcode Nextera indices (Detailed protocol: https://protocols.io/view/artic-neb-tagmentation-protocol-high-throughput-wh-bt66nrhe). Final libraries were pooled at equal volumes and cleaned at 0.7x (SPRI: Sample) using SPRIselect beads. Library was sequenced on Illumina Novaseq SP platform in a paired-end 2 × 150 cycle run.An incubation step with 1:10 dilution of FastSelect (Qiagen) reagent was included between the RNA fragmentation and first strand synthesis steps of the library prep to deplete highly abundant host rRNA sequences present in each sample. Equimolar pools (n=160-384 samples) of the resulting individual dual-barcoded library preps were subjected to paired-end 2 × 150bp sequence analysis on an Illumina NovaSeq 6000 (S2 or equivalent flow cell) to yield approximately 50 million reads per sample.

### Viral and Metagenomic Genome Alignment

For SARS-CoV-2 genomes, FASTQ sequences were aligned to the SARS-CoV-2 reference genome NC_045512.2 using minimap2.^36^ Non-SARS-CoV-2 reads were filtered out with Kraken2,^37^ using an index of human and viral genomes in RefSeq (index downloaded from https://genexa.ch/sars2-bioinformatics-resources/). Spiked primers for viral enrichment were trimmed from the ends of short reads using ivar.^38^ Finally, a pileup of the aligned reads was generated with samtools^39^, and consensus genomes were called with ivar. The full pipeline used is publicly available on Github (https://github.com/czbiohub/sc2-illumina-pipeline). All viral consensus sequences were uploaded to the GISAID database (https://www.gisaid.org/).

Host and metagenomic RNA alignment was performed using STAR run against a combined index of the human reference genome grch38, SARS-CoV2 (SARSCoV2_NC_045512.2), and ERCC spike-ins. STAR parameters were chosen to avoid bias towards GTAG eukaryotic splice signatures for both the viral RNA and host RNA analyses. Metagenomic classification of reads unmapped to both SARS-CoV2 and human was performed using KrakenUniq (https://genomebiology.biomedcentral.com/articles/10.1186/s13059-018-1568-0). KrakenUniq parameters (>=100 kmers and duplication <=less kmers) were chosen to avoid false positives. From the filtered KrakenUniq output, an abundance table was created by finding the kmer percentages (kmers divided by the total kmer count) for relevant taxa detected for each individual. This table included only well-represented taxa, which was defined as those appearing in at least 10% of patients. A uniform manifold approximation (UMAP) plot was then created from this table using fifteen nearest neighbors.

### Host Genome Sequence Alignment

Low-coverage FASTQ sequences underwent quality control assessment via FastQC v0.11.8 before alt-aware alignment to GRCh38.p12 using BWA-MEM v0.7.17-r1188. Duplicate sequences were marked with MarkDuplicates of the Picard Tools suite v2.21.2. After duplicate marking, base quality score recalibration was performed with Picard Tools’ BaseRecalibrator and high-confidence variant call sets from dbSNP and the 1000 Genomes Project. Quality control metrics, including coverage, were generated with Qualimap BAMQC v2.2.1, Samtools v1.10, and Mosdepth v0.2.9. Finally, quality control reports for each sample were aggregated using MultiQC v1.9. Reproducible code and steps are available at Protocols.io doi: (https://www.protocols.io/private/8CFBD1AD8FE611EA815E0A58A9FEAC2A) All high confidence calls were contributed to the COVID19 Host Genetics Initiative.^3^

### Variant Calling, Imputation, PCA, Kinshiship

BAM files were used for an initial calling with bcftools v1.9 mpileup.^40^ To account for the low-coverage sequencing we used the GLIMPSE algorithm v1.0 for imputation and phasing.^12^ Briefly, this algorithm uses a reference set of haplotypes (1000 genomes project samples in our case) to compute genotype likelihoods using a Gibbs sampling procedure. The imputed data were filtered for low imputation scores (INFO>0.8), and were then merged with a reference set that contained samples from: (1) the 1000 genomes project, (2) the Human Genome Diversity Project (HGDP),^41^ and (3) the Simons Genome Diversity Project (SGDP).^42^ While merging these data, we set minor allele count thresholds of 5 for our data and 20 for the reference set (e.g., MAC>4 using bcftools), and a stringent call rate threshold (*--geno 0*.*01* in PLINK2). ^43,44^ The resulting VCF was loaded into PLINK2 v2.00a3LM using the following flags: *dosage=DS, --import-dosage-certainty 0*.*8*. These merged data had 4,111,339 autosomal variants that survived the filters above. PLINK2 was then used for LD pruning (*--indep-pairwise 500 10 0*.*1*) and PCA (*--maf 0*.*01 --pca*). We also extracted the kinship matrix of our samples using the King algorithm (*--make-king* in PLINK2).^45^

### Genetic Ancestry Inference and assignation

Genetic ancestry was determined by running supervised local ancestry inference (RFMix v2.03)^46^ on the phased patient genomes using a training reference panel of single ancestry samples selected from the 1000 genomes project, HGDP, and SGDP via unsupervised genetic clustering (ADMIXTURE)^47^ at K = 7. Only individuals with greater than 0.95 assignment to one of the seven unsupervised clusters in this analysis were used as references for RFMix. The unsupervised cluster labels--African, East Asian, South Asian, Oceanian, European, West Asian, and Indigenous American--were chosen to reflect the biogeographic origin of the reference samples assigned to each cluster. Local genetic ancestry assignments along the genome were then summed to create overall genetic ancestry proportions for each sample which were used for barplots, covariates for regression analyses, and for making individual genetic ancestry assignments. Individual genetic ancestry labels (e.g. for determination of enrichment in cases vs. controls (Figure 2A), association with viral clades (Figure 2C) and controlling HLA associations with severe disease by genetic ancestry) were assigned based on these overall proportions via the following decision sequence: some Oceanian ancestry (Pacific Islanders) > 5%, some Indigenous American ancestry > 10%, West Asian > 50%, South Asian > 50%, East Asian > 50%, European > 50%, African > 50%. For individuals meeting none of these criteria, and ancestry label was assigned consisting of the two predominant ancestries (e.g. East Asian and European in Figure 2C).

### Association Analyses

Admixture mapping association analyses were used to regress the residual of severity of COVID symptoms for each patient--after correcting for associations with overall genetic ancestry proportion, BMI, sex, and age--against the local ancestry of each particular window of the genome for that patient.^48^ With the genome subdivided into 19,474 windows for local genomic ancestry assignment, and assuming complete independence between each, a naive Bonferroni corrected p-value of 2.57*10^-6 is obtained for genome-wide significance at p=.05; however, the genomic ancestry of neighboring, linked genomic windows is not independent and depends upon the characteristic length of each ancestry segment distribution, itself a function of the time since admixture in each population. A less stringent multiple-test correction factor that incorporates this distribution was empirically determined by considering the spectral density evaluated at frequency zero of an autoregressive model of local ancestries^17^, yielding an effective number of tests for each ancestry. This overall effective number of tests was taken over only samples that had at least 5% of that ancestry represented across their autosomes. Using this framework, together with the spectrum0.ar function implemented in the R package coda v 0.19, p-value thresholds for genome-wide significance at p=.05 for each ancestry were determined: African 2.54*10^-3, East Asian 3.89*10^-3, Indigenous American 1.15*10^-3, Oceanian 6.93*10^-3, South Asian 2.21*10^-3, and European/West Asian 1.83*10^-3.

### Host HLA Sequencing and Typing

Host genomic DNA samples ranging from 22-75ng were batched in sets of 46 plus one positive and one negative control. AllType™ FASTplex™ NGS Assay kits (One Lambda, A Thermo Fisher Scientific Brand, Canoga Park, CA) were used to prepare DNA sequencing libraries for 11 classical HLA genes (*HLA-A, HLA-C, HLA-B, HLA-DRB3, HLA-DRB4, HLA-DRB5, HLA-DRB1, HLA-DQA1, HLA-DQB1, HLA-DPA1*, and *HLA-DPB1*). As the success of the DNA sequencing is dependent on the initial target amplification and the subsequent library preparation, the following changes were made to the manufacturer’s protocol: 1) increased input DNA volume to 8.6 µl while maintaining the manufacturer’s recommended multiplex PCR protocol per sample; 2) eluted DNA in 12 µl of suspension buffer after the initial amplicon purification, and proceeded to the library preparation without normalization process; 3) increased the number of thermal cycles to 17 in the final DNA library amplification; 4) eluted DNA fragments in 22 µl for DNA sequencing. 500 µl of 1.3 pM DNA sequencing library was loaded into a MiniSeq Mid Output Kit (300-cycles) (FC-420-1004), and sequenced using MiniSeq DNA sequencer (Illumina Inc., San Diego, CA).

A total of 429 subjects (301 cases, 128 SARS-CoV-2 negative controls) yielded interpretable sequence reads to generate HLA genotypes. We supplemented these samples with sequencing of buffy coat or whole blood collected from high severity COVID19 patients collected from hospitalized patients (n=193). Fastq files were automatically imported into the TypeStream Visual NGS Analysis Software Version 2.0 upon the completion of DNA sequencing, and bioinformatically processed for DNA sequence assembly and HLA genotype assignments with IPD IMGT/HLA Database release version 3.39.0.^49^ We modified the software setting so that a maximum of 1.5 million sequences or 750,000 paired-end sequences are used for the sequence assembly and HLA allele assignments. We visually inspected the HLA genotype calls by the software, and made corrections as needed. The approved HLA genotype results were exported in Histoimmunogenetics Markup Language (HML) format,^50^ and generated comma separated value (CSV) reports for HLA genotypes, HLA serotypes including Bw4 and Bw6, KIR ligands (C1 and C2) and imputed HLA haplotypes. ^51,52^

Subjects were grouped in three categories (Negative: SS_MAX = 0; Mild: SS_MAX = 1 - 3; Severe: SS_MAX = 4 -8), and organized in six broad ancestry groups [European (EUR), Hispanic (HIS), Asian (ASI), African American (AFA), Native American (NAM) and Native Hawaiian/Pacific Islander (HPI)] based on self-reported ethnicity in clinical records. When self-reported ethnicity was not available, genetic ancestry calculated from the low pass WGS in this study was used as described above. We converted the genetic ancestry information to self-reported medical record ethnicity format as follows: European and West Asia => EUR; some Indigenous American and Hispanic => HIS; East Asian and South Asian = ASI; African => AFA; fully Indigenous American => NAM; Oceanian => HPI. We compared the distribution of both HLA serotypes and alleles from COVID19 positive individuals with low disease severity (maximum severity score 1-3, n=336) to those with high disease severity (maximum severity score 4-8, n=94). HLA serotype and allele frequencies were calculated in both Mild and Severe groups, and Odd Ratio (OR: Mild vs. Severe) and *p*-values were calculated for each serotype and allele using Bridging ImmunoGenomic Data-Analysis Workflow Gaps (BIGDAWG).^53^ Cochran-Mantel-Haenszel (CMH) tests^54^ were subsequently performed for all observed HLA-A, -B, -C, -DRB1, -DQB1 and -DPB1 serotypes and alleles across three major ethnic groups (EUR, HIS and ASI) using the “mantelhaen.test” function in stats R package. Subjects with AFA, NAM and HPI ethnic groups were excluded from CMH tests, because we had only 6, 1 and 5 subjects, respectively, that yielded HLA genotypes.

### Phylodynamic Analysis

For Bayesian inference of the viral phylogeny, we assumed the Extended Bayesian Skyline Plot ^55^ prior on the effective population size and coalescent prior on the phylogeny, a fixed molecular clock with a uniform prior distribution centered at 8×10^−4^ substitutions per site per year as done in ^56^. We assumed the HKY mutation model ^57^ with default hyperparameter priors in the BEAST2 software ^58^. We ran a Markov chain Monte Carlo chain to approximate the posterior distribution of the model parameters for 20 million iterations and thinned every 5000 iterations. The first 10% of samples were discarded as burn-in. We used Tracer ^59^ to assess the convergence and confirm that the effective sample size (ESS) was >120 for all parameters (except in 15% of effective population size parameters, estimations not shown). Finally, we used TreeAnnotator ^60^ to summarize the phylogeny posterior distribution and generated the maximum clade credibility tree of Figure S3. To test the association between clade composition and binary traits, we used the R package treeSeg.^61^

## Data Availability

We have established a resource of summary statistics for COVID19 severity versus host genotype by genetic ancestry, host HLA type, and metagenomic alignments (SEE LINK BELOW), and have contributed host genetic summary data and viral consensus sequences to the COVID19 Host Genetics Initiative(1) and GISAID (GISAID.org,2) respectively.
(1) Initiative, C.19 H. G. & COVID19 Host Genetics Initiative. Mapping the human genetic architecture of COVID19. Nature (2021)
(2) Elbe, S. & Buckland-Merrett, G. Data, disease and diplomacy: GISAIDs innovative contribution to global health. Glob Chall 1,(2017).

https://covid-omics.org/results

## Supplementary Material

### Supplementary Text

#### Additional detail regarding HLA-Severity associations

The *HLA-C*15:02* allele is common in some Native American tribes from North, Central and South America. The alleles of *HLA-C* can be classified into the C2 and C1 groups according their interaction with the inhibitory receptors KIR2DL1 and KIR2DL2/3 present in NK cells, respectively; the allele *HLA-C*15:02* belongs to the C2 group. The current study found no significant associations of other *HLA-C* alleles of the C2 group or of other HLA ligands of KIR receptors suggesting that the broad KIR/HLA ligand interactions are not major determinants of disease course. We could not confirm the risk reduction in the prototype allele *HLA-B*15:01* for the serotype B62, because we observed other B62 alleles (*HLA-B*15:04, -B*15:25, -B*15:27, -B*15:35, -B*15:48* and *-B*15:146*). They are mostly HLA alleles for Hispanic and Asian. The *HLA-B*15:01* allele was recently associated with asymptomatic SARS-CoV-2 infection in European ancestry subjects. In our study subjects only 32% percent of the subjects had European ancestry; we did not find statistical significance for the individual allele *HLA-B*15:01* that is by far more common than other alleles of the serotype B62 in Caucasians. The alleles *HLA-B*15:04*, -*B*15:25, -B*15:27, -B*15:35, -B*15:48* and *-B*15:146* are found in subjects with Hispanic and Asian. The B*15 alleles corresponding to the B62 serotype detected in the present study show high structural homology (differing by 1-5 amino acid replacements) and share identity in the B-pocket substructure that determines what type of peptide residue at the second position is anchored. A recent study in outcomes of hematopoietic stem cell transplantation found that the HLA-B62 supertype was associated with decreased transplant-related mortality in the entire patient cohort.^62^ It can be speculated that HLA-B62 may play a role in immune response; the associations identified in the present study lend support to the hypothesis that peptide presentation by alleles of the B62 serotype in addition to the *HLA-C*15:02* allele may contribute to beneficial outcomes in SARS-CoV-2 Disease.

### Supplementary Tables

**Supplementary Table 1.** WHO-based COVID-19 Clinical Severity Scale

| Score | Patient Status | Description | EHR annotations |
| --- | --- | --- | --- |
| 0 | Uninfected | No clinical or virological evidence of infection | SARS-CoV-2 negative |
| 1 | Ambulatory | Asymptomatic | Non COVID-related symptoms |
| 2 |  | Symptomatic | COVID related symptoms (cough, fever, shortness of breath, chest pain, tachycardia, acute pharyngitis, fever, others) |
| 3 | Hospitalized, Mild disease | Hospitalized, no oxygen | COVID related symptoms + hospitalization time (without oxygen support) |
| 4 |  | Hospitalized, oxygen mask | Nasal cannula; face mask; high-oxygen mask; aerosol mask; tracheal collar; others |
| 5 | Critical Care, Severe disease | non-invasive ventilation / high flow oxygen | High-flow nasal cannula; non-invasive positive pressure ventilation |
| 6 |  | intubation and mechanical ventilation | Invasive Ventilation; endotracheal tube |
| 7 |  | intubation + organ support | Intubation + dobutamine, dopamine, epinephrine, norepinephrine, phenylephrine, vasopressin |
| 8 | Dead | Deceased | Deceased |

### Supplementary Figures

**Figure S1.**
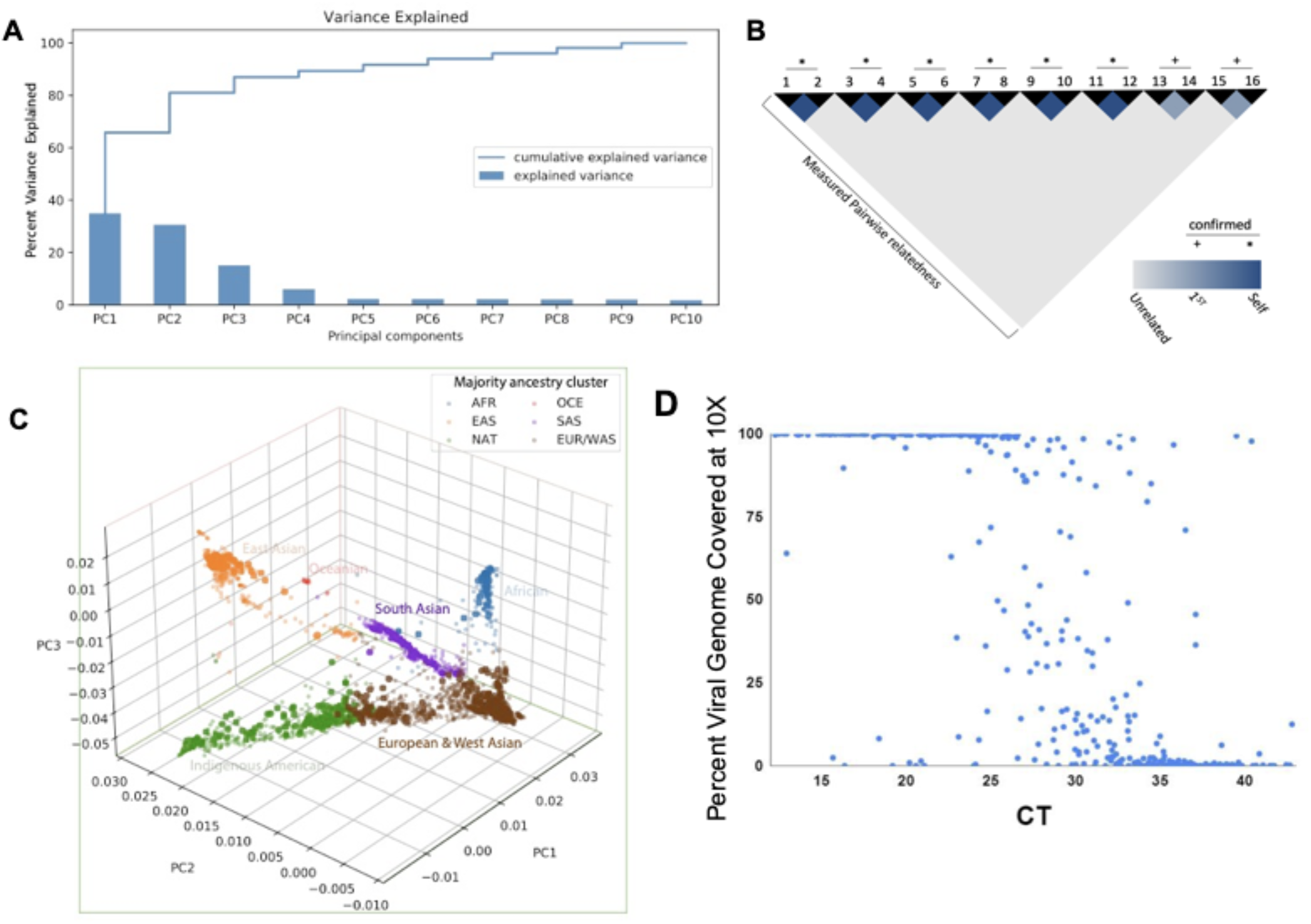
**(A)** Variance explained by the top 10 PCs. The first four PCs explain ∼90% of variance. **(C)** Genomic data of patients (bold points) projected onto top three PCs created by samples from 1000 Genomes, HGDP and SGDP (light points). **(B)** Genotyping by low pass WGS recapitulates known kinship relationships. **(D)** Viral genome recovery using ARTIC primer capture was improved at higher CT values compared with shotgun RNAseq (Figure 1D).

**Figure S2.**
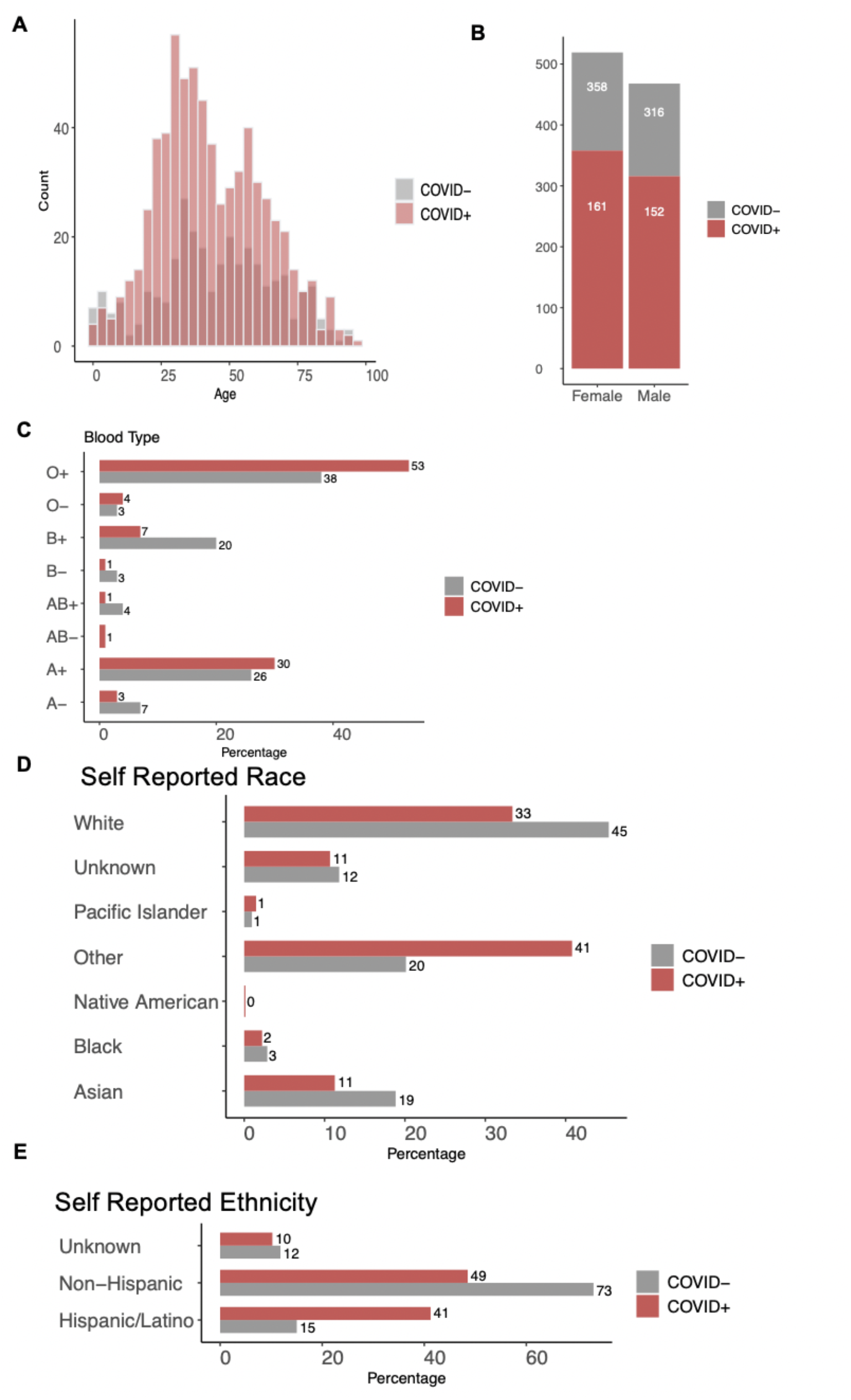
Clinical Characteristics of COVID19 positive individuals versus negative controls.

**Figure S3.**
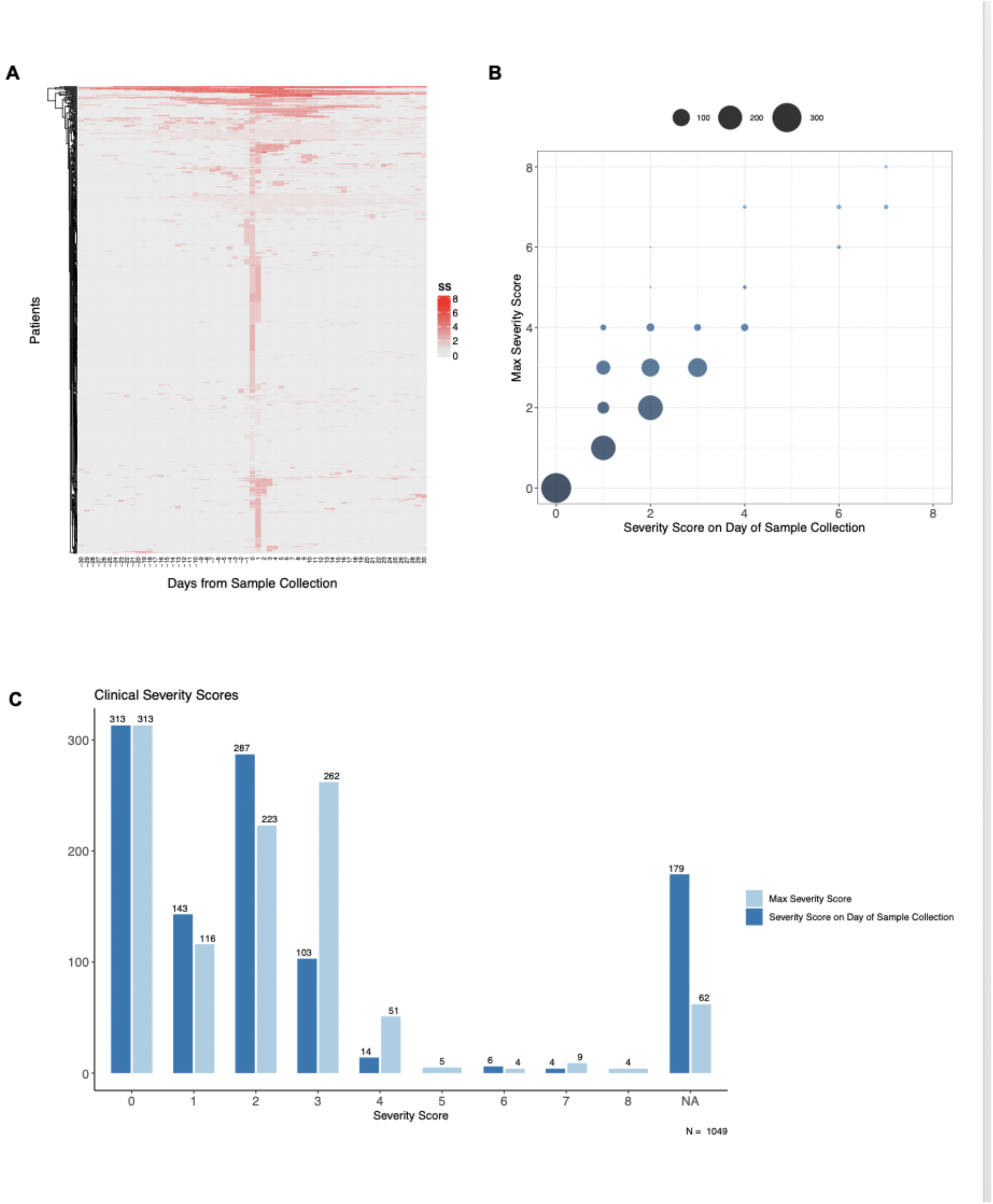
Severity score. (A) Variance in severity score for 30 days before and after NP swab collection for all samples. (B) Correlation between severity on day of sample collection and highest severity (Max Severity Score) in the 30 days before and after swab collection. (C) Redistribution of severity scores in the 30 days before and after collection vs day of collection.

